# Detection of cognitive decline using a single-channel EEG with an interactive assessment tool

**DOI:** 10.1101/2020.08.13.20171876

**Authors:** Lior Molcho, Neta B. Maimon, Narkiss Pressburger, Noa Regev-Plotnik, Sarit Rabinowicz, Nathan Intrator, Ady Sasson

## Abstract

**Background:** Cognitive decline remains highly underdiagnosed despite efforts to find novel biomarkers for detection. EEG biomarkers based on machine learning may offer a noninvasive low-coast approach for identifying cognitive decline. However, most studies use multi-electrode systems which are less accessible. This study aims to evaluate the ability to extract cognitive decline biomarkers using a wearable single-channel EEG system with an interactive assessment tool.

**Methods:** This pilot study included data collection from 82 participants who performed a cognitive assessment while being recorded with a single-channel EEG system. Seniors in different clinical stages of cognitive decline (healthy to mild dementia) and young healthy participants were included. Seniors’ MMSE scores were used to allocate groups with cutoff scores of 24 and 27. Data analysis included correlation analysis as well as linear mixed model analysis with several EEG variables including frequency bands and three novel cognitive biomarkers previously extracted from a different dataset.

**Results:** MMSE scores correlated significantly with reaction times, as well as two EEG biomarkers: A0 and ST4. Both biomarkers showed significant separation between study groups: ST4 separated between the healthy senior group and the low-MMSE group. A0 differentiated between the healthy senior group and the other three groups, showing different cognitive patterns between different stages of cognitive decline as well as different patterns between young and senior healthy participants. In the healthy young group, activity of Theta, Delta, A0 and VC9 biomarkers significantly separated between high and low levels of cognitive load, consistent with previous reports. VC9 and Theta showed a finer separation between low cognitive load levels and resting state.

**Conclusions:** This study successfully demonstrated the ability to assess cognitive states with an easy-to-use portable single-channel EEG device with an interactive cognitive assessment. The short set-up time and novel biomarkers enable objective and easy assessment of cognitive decline. Future studies should explore potential usefulness of this tool in characterizing changes in EEG patterns of cognitive decline over time, for detection of cognitive decline on a large scale in every clinic to potentially allow early intervention.

## Background

Cognitive decline is characterized by impairments in various cognitive functions such as memory, orientation, language, and executive functions [1], expressed more than anticipated for an individual’s age and education level. Cognitive decline with memory deficit indications is associated with high-risk for developing dementia in general, and Alzheimer’s disease in particular [2]. Interventions starting early in the disease process, before substantial neurodegeneration has taken place, can change the progression of the disease dramatically [3]. Yet, there is still no universally recommended screening tool that satisfies all needs for early detection of cognitive decline [4].

The most commonly used screening tool for cognitive decline in the elderly population is the Mini-Mental State Examination (MMSE) [5]. The MMSE evaluates cognitive function, producing a total possible score of 30 points. Patients who score below 24 would typically be suspected of cognitive decline or early dementia [6]. However, several studies showed that factors unrelated to the cognitive state, such as age and education as well as tester bias, could affect individual scores [7], [8].

Naturally, objective cognitive assessment based on brain activity measurements would be preferable to subjective clinical evaluation using pen-and-paper assessment tools like the MMSE. Electroencephalography (EEG) offer such noninvasive and relatively inexpensive screening tool for cognitive assessment [9]. EEG studies investigating cognitive decline highlight the role of theta power as a possible indicator for early detection of cognitive decline [10]–[12]. Specifically, it was found that frontal theta activity differs substantially in cognitively impaired subjects performing cognitive tasks compared to healthy seniors. Studies suggest that novel diagnostic classification based on EEG signals could be even more useful than frontal theta for differentiating between clinical stages [10]. The development of machine learning (ML) alongside high-level signal processing, has largely contributed to the extraction of useful information from the raw EEG signal [13]. Novel techniques are capable of exploiting the large amount of information on time-frequency processes in a single recording [14], [15]. Recent studies demonstrated novel measures of EEG for identification of cognitive impairment with high accuracy, using classifiers based on neural networks, wavelets, and blind source separation, indicating the relevance of such methods for cognitive assessment [16]–[19].

However, most studies in this field have several constraints. Most commonly, such studies use multichannel EEG systems to characterize cognitive decline. The difficulty with multichannel EEG is the long set-up time which require specifically trained technicians, as well as a professional interpretation of the results. This makes the systems costly and not portable, thus, not suitable for wide-range screening in every clinic. Consequently, these are not included in the usual clinical protocol for cognitive decline detection. This emphasizes the need for additional cost-effective tools with a short assessment time and easy set-up, to allow detection of cognitive decline of patients in the community.

A recent study [20] examined differences in responses to auditory stimuli between cognitively impaired and healthy subjects and concluded that cognitive decline can be characterized using data from a single EEG channel. Specifically, using data from frontal electrodes, the authors extracted 590 features that were later used in classification models to identify subjects with cognitive impairments. The results contribute to the idea that a single channel EEG can be used as an efficient and convenient way for detecting cognitive decline. However, the risk of overfitting the data in such classification studies should be addressed to ensure generalization capabilities, especially with a small sample size. Studies that use the same dataset for training as well as feature extraction [20]–[22], extend the risk of overfitting. For generalization of the data, the features should be examined on different datasets and provide consistency in the results on new datasets. Furthermore, measuring the correlations of the extracted features with standard clinical measurements (like the MMSE score) or behavioral results of cognitive tests (like reaction times and accuracy) is highly valuable for further validation of such novel features.

In this study we evaluated the ability of an easy-to-use single-channel EEG system with an interactive cognitive assessment to potentially detect cognitive decline in the elderly population. The system uses auditory stimuli, and extracts biomarkers using harmonic analysis and machine learning methods from the EEG signal. The pre-extracted biomarkers used in this study were validated in a previous study performed on young healthy subjects [23]. This pilot study aims to evaluate the ability of the system to extract cognitive decline biomarkers, recognizing the importance of providing an accurate low-cost alternative for cognitive decline detection.

## Methods

### Participants

#### Senior participants

Ethical approval for this study was granted by the Ethics Committee (EC) of Dorot Geriatric Medical Center on July 01, 2019. Israeli Ministry of Health (MOH) registry number MOH_2019-10-07_007352, first posted on Oct 07, 2019. NIH Clinical Trials Registry number NCT04386902, first posted on May 13, 2020, URL: https://clinicaltrials.gov/ct2/show/NCT04386902?term=Neurosteer&draw=2&rank=1.

Sixty patients from the inpatient rehabilitation department at Dorot Geriatric Medical Center were recruited to this study. For the full demographic details see Table 1. The overall mean age was 77.55 (9.67) years old. There was a wide range of ages for each group with no significant age difference between the groups. Participants consisted of 47% females and 53% males. Among the patients, 82% were hospitalized for orthopedic rehabilitation and 18% due to various other causes. Among the patients who had surgery, an average of 27 (16.3) days passed since the surgery. Potential subjects were identified by the clinical staff during their admission to the inpatient rehabilitation department. All subjects were hospitalized at the center and were chosen based on inclusion criteria specified in the study protocol. The patients undergo a Mini Mental State Examination (MMSE) by an Occupational Therapist upon hospital administration and this score was used for screening patients to include those that have scores between 10-30. All subjects were also evaluated for their ability to hear, read, and understand instructions for discussion of Informed Consent Form (ICF) as well as for the auditory task. Patients that speak English, Hebrew, and Russian were provided with the appropriate ICF and auditory task in the language they could read and understand. All participants provided ICF according to the guidelines outlined in the Declaration of Helsinki. Patients that showed any verbal or non-verbal form of objection were not included in the study. Other exclusion criteria included MMSE score lower than 10, presence of several comorbidities, damage to integrity of scalp and/or skull and skin irritation in the facial and forehead area, significant hearing impairments, and history of drug abuse.

**Table 1.**
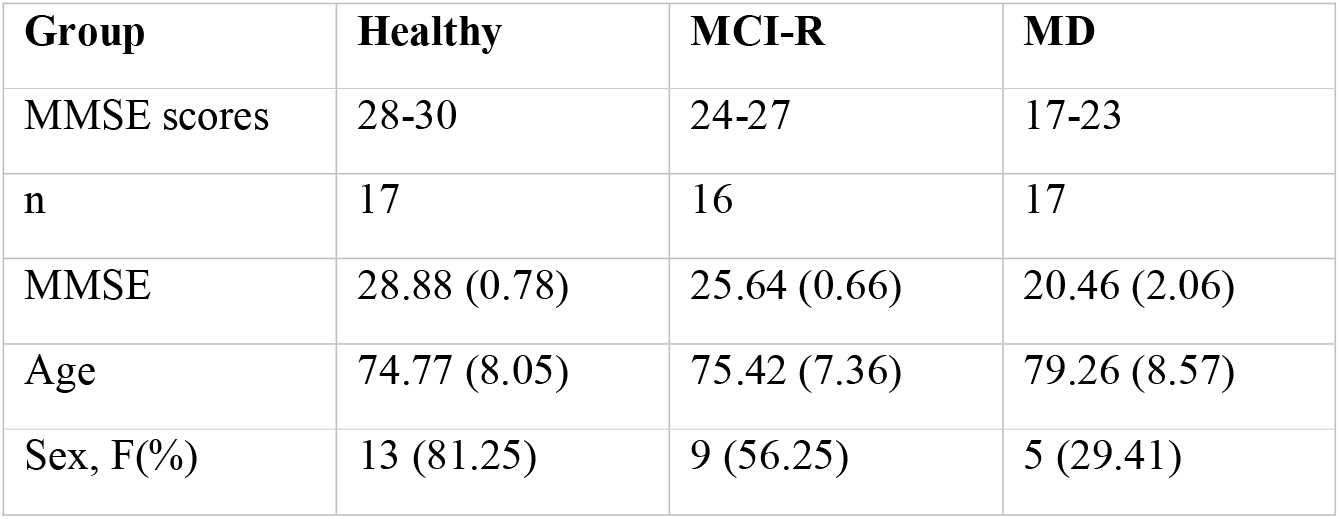
Demographic information of the senior groups included in the analysis.

| **Group** | **Healthy** | **MCI-R** | **MD** |
| --- | --- | --- | --- |
| MMSE scores | 28-30 | 24-27 | 17-23 |
| n | 17 | 16 | 17 |
| MMSE | 28.88 (0.78) | 25.64 (0.66) | 20.46 (2.06) |
| Age | 74.77 (8.05) | 75.42 (7.36) | 79.26 (8.57) |
| Sex, F(%) | 13 (81.25) | 9 (56.25) | 5 (29.41) |

In total, 50 of the 60 recruited patients, completed the auditory task and their EEG data was used. 10 patients signed the ICF and were included in the overall patient count but were excluded from data analysis due to their desire to stop the study or technical problems in the recording. The participants were divided into 3 groups according to the MMSE scores: 17 patients with a score of 17-23 in the Mild Dementia (MD) group, 16 patients with a score of 24-27 in the Mild Cognitive Impairment Risk (MCI-R) group and 17 patients with a score of 28-30 in the Healthy seniors group. See table 1 for demographic information.

#### Healthy young participants

22 healthy students participated in this study for course credit. The overall mean age was 24.09 (2.79) years old. Participants consisted of 60% females and 40% males. Ethical approval for this study was granted by Tel-Aviv University ethical committee 27.3.18.

### Apparatus

#### EEG device

EEG recordings were performed using a single channel EEG system (Aurora Neurosteer Inc). A 3-electrode medical-grade patch was placed on the subject’s forehead using dry gel for optimal signal transduction. The electrodes were located at Fp1 and Fp2 and a reference electrode at Fpz. EEG signal was amplified by a factor of a 100 and sampled at 500 Hz. Signal processing was done in the Neurosteer cloud, for further details see Appendix A.

#### EEG Recording and Auditory battery

The recording room was quiet and illuminated. The research assistant set up the sanitized system equipment (electrode patch, sensor, EEG monitor, clicker) and provided general instructions to the participants before starting the task. Then the electrode was placed on the subject’s forehead and the recording was initiated. The participant was sitting during the assessment and heard instructions through a loudspeaker connected to the EEG monitor. The entire recording session typically lasted 20-30 minutes. The cognitive assessment battery was pre-recorded and included a detection task as well as answering a series of true/false questions by pressing on a wireless clicker button. Further explanations for the task were kept at a minimum to avoid bias. A few minutes of baseline activity were recorded to ensure an accurate test. The auditory cognitive assessment lasted 18 minutes.

#### Detection Task

Figure 1 illustrates the detection task used in the study. In each block, participants were presented with a sequence of melodies (played by a violin, a trumpet, and a flute). The participants were given a clicker to respond to the stimuli. In the beginning of each block, auditory instructions indicated an instrument to which the participant responded by clicking once. The click response was only to “yes” trials, when the indicated instrument melody had played. The task included two difficulty levels to test increasing cognitive load. In level 1, each melody was played for 3 seconds, and the same melody repeated throughout the entire block. The participant was asked to click once as fast as possible for each repetition of the melody. In level 2, the melodies were played for 1.5 seconds, and all three instruments appeared in the block. The participants were asked to click only for a specific instrument within the block and ignore the rest of the melodies.

**Figure 1.**
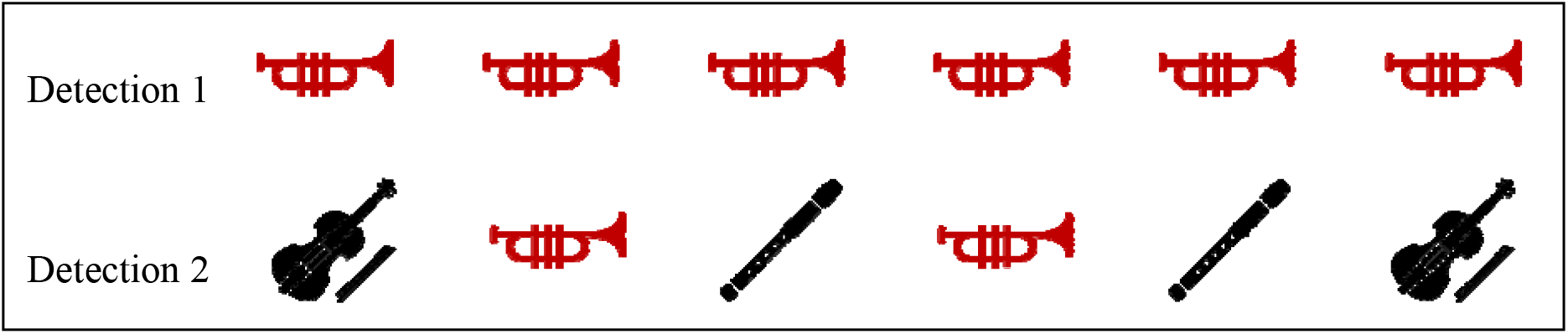
An example of six trials of detection level 1 (top) and detection level 2 (bottom). Both examples show a “Trumpet block” in which the participant reacts to the trumpet melody. Red icons represent trials in which the participant was required to respond with a click when hearing the melody, indicating a “yes” response.

### Data Analysis

#### Construction of Brain Activity Features and Classifiers

The signal processing algorithm interprets the EEG data using a time/frequency wavelet-packet analysis [24], [25], which characterizes different components by their fundamental frequency and corresponding higher harmonics. In addition, standard spectral analysis of the signal was used to produce the classical EEG frequency bands.

The optimal number of different features was determined on a large dataset using unsupervised Machine Learning techniques (i.e. ML extracting features on unlabeled data). This analysis resulted in 121 Brain Activity Features (BAFs). The BAFs activity can be regarded as a series of vectors with 121 values that are sampled every second. Combinatorically, there are millions of linear and non-linear combinations that can be created out of these long vectors. Prior to this examination, several such combinations were computed using ML. These combinations were calculated from data of healthy subjects performing different tasks in different difficulty levels. The labelled data was used to train linear and non-linear classifiers that differentiate between different tasks. The value of each feature was calculated once every second from a moving window of 4 seconds. Three of those biomarkers were described in a previous study on healthy subjects performing a well-validated cognitive task (i.e. n-back task) [23]. The biomarkers exhibited separation between different levels of cognitive load and therefore were the most relevant for the present study. The full technical specifications regarding the construction of these features and the extractions of the biomarkers are provided in Appendix A.

### Dependent variables

#### Behavioral measurements

The behavioral dependent variables included mean response accuracy and mean reaction times (RT) per participant, for correct responses only.

#### Electrophysiological variables

The electrophysiological dependent variables included the power spectral density. Absolute power values were converted to logarithm base 10 to produce values in dB. The following frequency bands were included: Delta (0.5-4 Hz), Theta (4–7 Hz), Alpha (8–15 Hz), Beta (16–31 Hz), and lower Gamma (32–45 Hz).

The BAFs analysis included the activity of the three selected biomarkers: ST4, A0, and VC9 normalized to a scale of 0-100. The EEG variables were calculated per each second from a moving window of 4 seconds, and mean activity per condition was taken into the analyses.

#### Statistical analyses

Statistical analyses were performed on data from 50 senior participants and 22 young participants. Analyses included Pearson correlations and Mixed Linear Models (LMM).

#### Pearson correlations

Based on previous studies [3] we hypothesized that the Reaction Times (RTs) in the cognitive detection task would be higher for participants with lower MMSE scores. This was tested by calculating Pearson correlation between mean RTs in detection levels 1 and 2, and the individual MMSE score of each participant. RTs correlations to the EEG variables were also calculated.

Another correlation analysis was performed to find which of the EEG variables correlated with the (previously assigned) MMSE score of each participant. For this purpose, Pearson correlation was calculated using the mean activity of the EEG variables during the detection task and each individual MMSE score.

#### Linear Mixed Models (LMM)

The independent variables included task as a within-participants variable (including detection 1, detection 2, resting state tasks) and group as a between-participants variable. Groups were allocated as follows (see Figure 2): data of the senior participants was divided into three groups according to MMSE scores: Mild Dementia (MD) group with MMSE scores between 17-24, Mild Cognitive Impairment Risk (MCI-R) group with MMSE scores between 24-27, and Healthy Seniors with MMSE scores between 28-30. We used MMSE score cutoffs of 24 and 27 in allocating the groups as we are mostly interested in detecting cognitive decline as early as possible and found previous indications that a higher cutoff score would achieve optimal evaluations of diagnostic accuracy [7]. Furthermore, it was argued that educated individuals who score below 27 are at greater risk of being diagnosed with dementia [26]. The last group included in the analysis was consist of the 22 young healthy participants (Healthy young group). Overall, 4 groups were taken into the analysis. Since both independent variables have more than two levels, indicator variables (aka dummy variables) were computed. For the group variable, Healthy seniors group was determined as the reference level, and the three remaining levels were computed as dummy variables: Healthy young (e.g., the difference between Healthy seniors and Healthy young), MCI-R (e.g., the difference between Healthy seniors and MCI-R), and MD (e.g., the difference between Healthy seniors and MD). Accordingly, a significant effect in one of the groups would mean that the activity of the Healthy seniors group is significantly different than the activity of said group. For the task variable, detection 1 (which represents low cognitive load level) was determined as the reference level, and the two remaining levels were computed as dummy variables: Detection 2 (e.g., the difference between detection 1 and detection 2 which represents high cognitive load level) and Resting state (e.g., the difference between detection 1 and resting state). A significant effect of one of the levels, would mean that the activity during detection 1 level was significantly different than the activity during said level.

**Figure 2.**
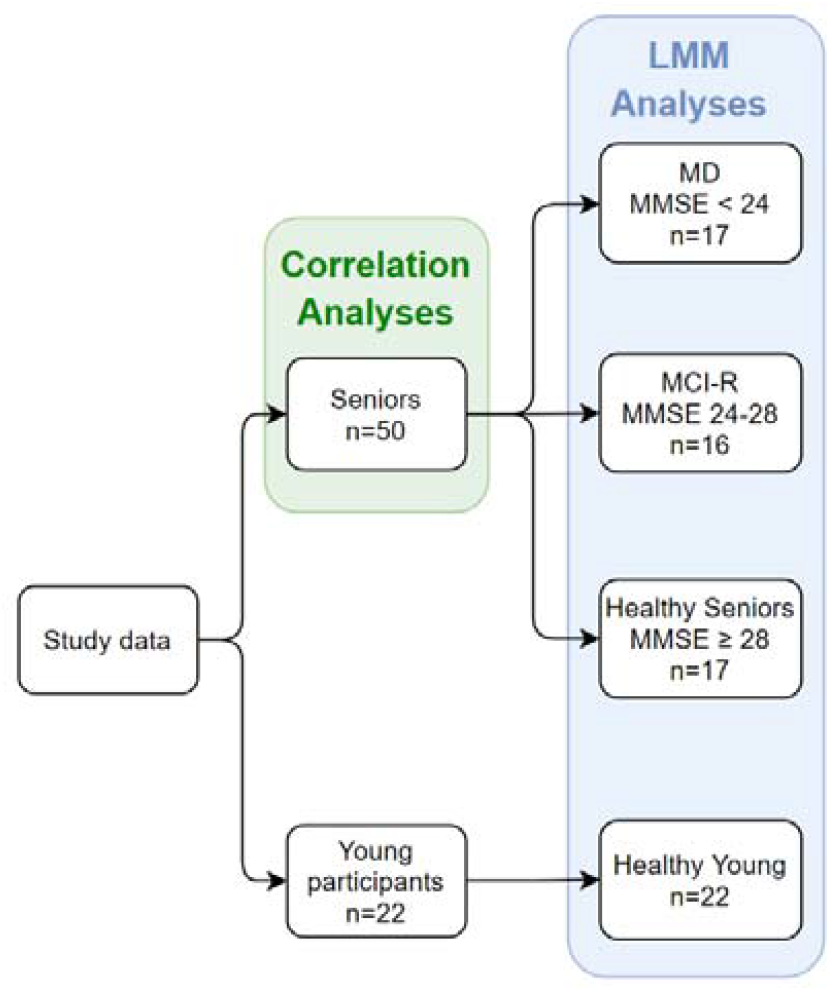
Study design and groups at each stage. The study included both seniors and young participants as controls. For the senior participants, an MMSE score was obtained, and division into groups was based on the individual MMSE score.

An LMM analysis was performed on each EEG variable separately (A0, ST4, VC9, Delta, Theta, Alpha, Beta, and Gamma). Due to the interpretation complexity of the effects, the first analysis included the two variables (group and task level) without their interactions. Significance level for this analysis was p<0.05. For models that achieved significant effects in the task variable, follow-up models were calculated to examine the activity level differences within each group. Therefore, the interaction between task level and group was examined within the senior groups and the young healthy group separately. For these post-hoc models (measuring the simple effects of task within each study group), significance level resulted in the LMM model was corrected with Bonferroni correction to p<0.025 (in accordance with two comparisons). all analyses were conducted with Python Statsmodels [27].

## Results

### Validation of behavioral task

Correlations between individual MMSE scores and participants’ Reaction Times (RTs) in both levels of the detection task were significant, both for each level separately as well as the mean activation in the entire cognitive detection task (*p<*.01 for all, see Table 2). Additionally, mean RTs were calculated for each participant and each task. A0 activity significantly increased with slower participant RTs, while ST4 activity significantly decreased with slower RTs (see Table 3 and Figure 3).

**Table 2.** Results of Pearson correlation between individual MMSE scores and reaction times (RTs) for the cognitive detection task (levels 1 and 2) and mean reaction times. Significant effects are marked in bold.

| Task level | Detection 1 | Detection 2 | Mean |
| --- | --- | --- | --- |
| r | -.535** | -.423** | -.519** |
| p | **<.001** | **0.005** | **<.001** |
| df | 49 | 43 | 49 |

**Table 3.** Pearson correlation analysis between individual mean RTs of the cognitive detection task (levels 1 and 2) and EEG variables activity. Significant effects are marked in bold.

| Feature | Alpha | Beta | Delta | Gamma | Theta | A0 | VC9 | ST4 |
| --- | --- | --- | --- | --- | --- | --- | --- | --- |
| r | 0.198 | 0.194 | 0.037 | 0.183 | 0.000 | 0.264 | -0.036 | -0.252 |
| p | 0.058 | 0.064 | 0.728 | 0.081 | 0.997 | **0.011** | 0.732 | **0.016** |

**Figure 3.**
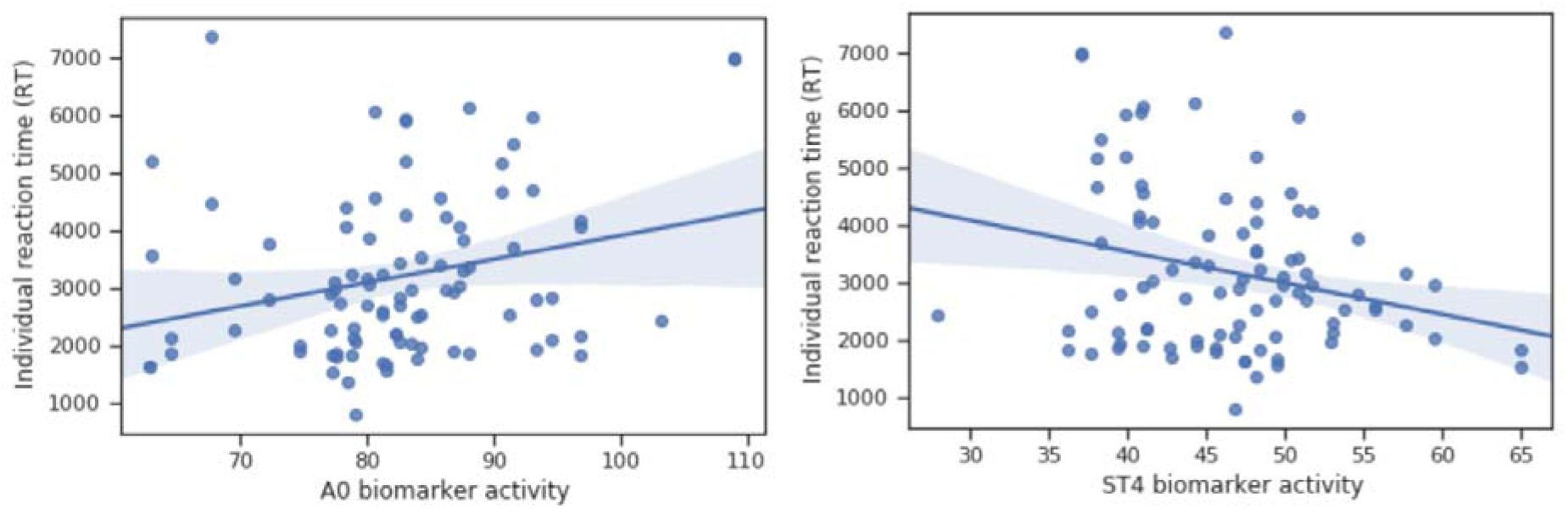
Pearson correlation between individual mean reaction times (RTs) and mean activity of A0 (left) and ST4 (right) during detection task.

### Correlations between MMSE scores and EEG variables

Pearson r and p values of correlations for each EEG variable are presented in Table 4. The activity of A0 increased for lower MMSE score while the activity of ST4 decreased for lower MMSE score (*p*=.010 and *p*=.009, respectively, see Figure 4). All other EEG variables did not exhibit significant correlation with MMSE scores.

**Table 4.** Pearson correlation analysis between individuals MMSE scores and mean activity of the EEG variables during the detection task. Significant effects are marked in bold.

| Feature | Alpha | Beta | Delta | Gamma | Theta | A0 | VC9 | ST4 |
| --- | --- | --- | --- | --- | --- | --- | --- | --- |
| r | -0.1111 | -0.2430 | 0.0773 | -0.2269 | 0.1439 | -0.3570 | 0.0930 | 0.3617 |
| p | 0.4423 | 0.0891 | 0.5938 | 0.1130 | 0.3187 | **0.0109** | 0.5204 | **0.0099** |

**Figure 4.**
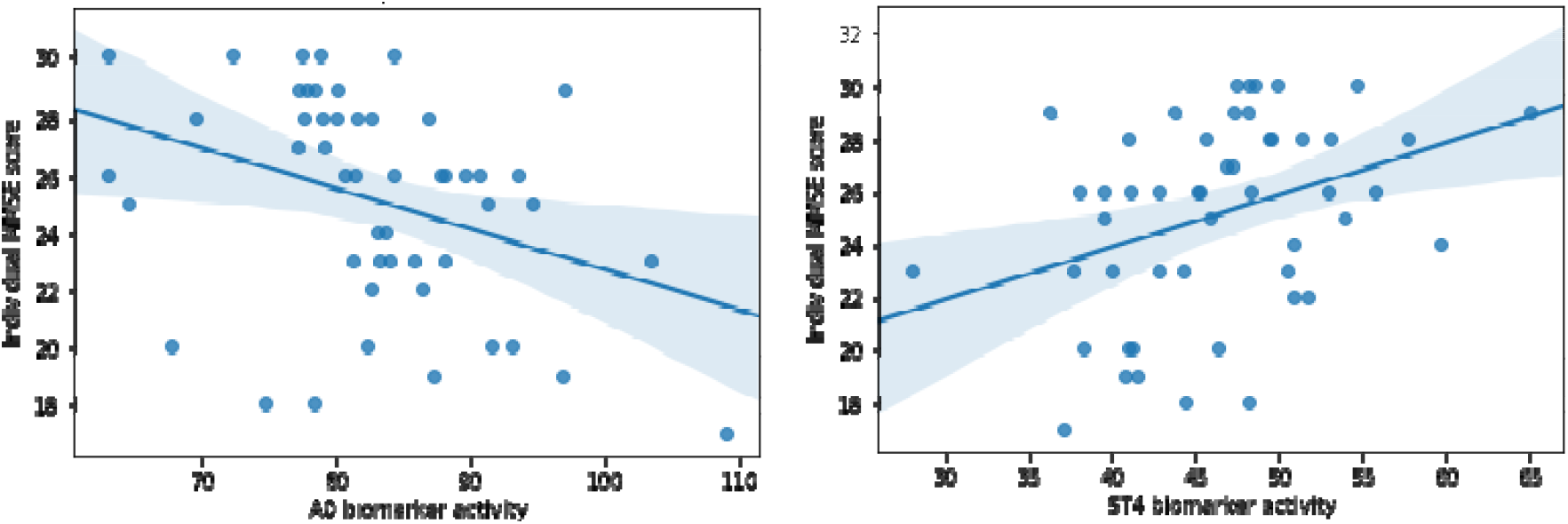
Pearson correlation between individual MMSE scores and mean activity of A0 (left) and ST4 (right) during detection task.

### Mixed linear models results

The first LMM analysis (see table 5), revealed significant differences between the groups in VC9, A0 and ST4 biomarkers’ activity. Specifically, VC9 activity was significantly lower in healthy young participants compared to healthy seniors (*p=*0.005), A0 showed significant differences between healthy seniors and all other groups (Healthy young, *p<*0.001, MCI-R, *p=*0.04 and MD *p=*0.011), and ST4 showed a significant difference between healthy seniors and MD group (*p=*0.011).

**Table 5.** Coefficients, Standard errors, z values, p values, and CI interval for group (healthy seniors as reference group), and task level (detection 1 as reference group) in the first LMM analysis which not included interactions.

| Feature | Variable [Level] | Coef. | Std.Err. | z | P>\|z\| | [0.025 | 0.975] |
| --- | --- | --- | --- | --- | --- | --- | --- |
| A0 | Intercept | 78.979 | 1.675 | 47.15 | <0.001 | 75.696 | 82.262 |
|  | Group [Healthy young] | -12.58 | 2.278 | -5.522 | **<0.001** | -17.046 | -8.115 |
|  | Group [MCI-R] | 4.867 | 2.367 | 2.056 | **0.04** | 0.227 | 9.507 |
|  | Group [MD] | 5.935 | 2.342 | 2.534 | **0.011** | 1.344 | 10.525 |
|  | Task Level [Detection 2] | 1.098 | 0.461 | 2.383 | **0.017** | 0.195 | 2.002 |
|  | Task Level [Resing State] | 0.648 | 0.549 | 1.181 | 0.238 | -0.428 | 1.723 |
| ST4 | Intercept | 47.912 | 1.129 | 42.448 | <0.001 | 45.7 | 50.125 |
|  | Group [Healthy young] | 0.775 | 1.4 | 0.554 | 0.58 | -1.969 | 3.52 |
|  | Group [MCI-R] | -1.231 | 1.555 | -0.792 | 0.428 | -4.279 | 1.816 |
|  | Group [MD] | -3.829 | 1.512 | -2.532 | **0.011** | -6.792 | -0.865 |
|  | Task Level [Detection 2] | 0.38 | 0.383 | 0.992 | 0.321 | -0.37 | 1.13 |
|  | Task Level [Resing State] | -0.597 | 0.636 | -0.938 | 0.348 | -1.844 | 0.65 |
| VC9 | Intercept | 57.243 | 1.239 | 46.183 | <0.001 | 54.814 | 59.672 |
|  | Group [Healthy young] | -4.695 | 1.687 | -2.784 | **0.005** | -8.001 | -1.39 |
|  | Group [MCI-R] | -0.145 | 1.729 | -0.084 | 0.933 | -3.533 | 3.243 |
|  | Group [MD] | -1.487 | 1.707 | -0.871 | 0.384 | -4.832 | 1.858 |
|  | Task Level [Detection 2] | 1.174 | 0.322 | 3.647 | **<0.001** | 0.543 | 1.805 |
|  | Task Level [Resing State] | -1.611 | 0.476 | -3.384 | **0.001** | -2.544 | -0.678 |
| Theta | Intercept | -4.864 | 0.731 | -6.657 | <0.001 | -6.296 | -3.432 |
|  | Group [Healthy young] | -1.838 | 1.006 | -1.826 | 0.068 | -3.811 | 0.135 |
|  | Group [MCI-R] | -0.565 | 1.026 | -0.551 | 0.582 | -2.575 | 1.445 |
|  | Group [MD] | -1.512 | 1.006 | -1.504 | 0.133 | -3.483 | 0.458 |
|  | Task Level [Detection 2] | 0.561 | 0.175 | 3.209 | **0.001** | 0.219 | 0.904 |
|  | Task Level [Resing State] | -0.941 | 0.29 | -3.249 | **0.001** | -1.508 | -0.373 |
| Delta | Intercept | 4.743 | 0.855 | 5.547 | <0.001 | 3.067 | 6.418 |
|  | Group [Healthy young] | 0.597 | 1.156 | 0.516 | 0.606 | -1.669 | 2.863 |
|  | Group [MCI-R] | 0.423 | 1.237 | 0.342 | 0.733 | -2.002 | 2.847 |
|  | Group [MD] | -0.866 | 1.189 | -0.729 | 0.466 | -3.196 | 1.463 |
|  | Task Level [Detection 2] | 0.667 | 0.245 | 2.723 | **0.006** | 0.187 | 1.148 |
|  | Task Level [Resing State] | -0.645 | 0.357 | -1.806 | 0.071 | -1.344 | 0.055 |
| Alpha | Intercept | -11.099 | 0.519 | -21.398 | <0.001 | -12.115 | -10.082 |
|  | Group [Healthy young] | 1.053 | 0.711 | 1.481 | 0.139 | -0.34 | 2.447 |
|  | Group [MCI-R] | 0.674 | 0.735 | 0.916 | 0.36 | -0.768 | 2.115 |
|  | Group [MD] | -0.242 | 0.718 | -0.337 | 0.736 | -1.648 | 1.165 |
|  | Task Level [Detection 2] | 0.212 | 0.121 | 1.756 | 0.079 | -0.025 | 0.448 |
|  | Task Level [Resing State] | -0.139 | 0.183 | -0.76 | 0.447 | -0.499 | 0.22 |
| Beta | Intercept | -10.479 | 0.888 | -11.797 | <0.001 | -12.22 | -8.738 |
|  | Group [Healthy young] | 1.156 | 1.169 | 0.988 | 0.323 | -1.136 | 3.447 |
|  | Group [MCI-R] | 1.538 | 1.263 | 1.218 | 0.223 | -0.938 | 4.013 |
|  | Group [MD] | 1.343 | 1.244 | 1.08 | 0.28 | -1.095 | 3.782 |
|  | Task Level [Detection 2] | 0.259 | 0.189 | 1.369 | 0.171 | -0.112 | 0.629 |
|  | Task Level [Resing State] | 0.467 | 0.267 | 1.751 | 0.08 | -0.056 | 0.99 |
| Gamma | Intercept | -11.006 | 0.961 | -11.455 | <0.001 | -12.889 | -9.123 |
|  | Group [Healthy young] | -0.156 | 1.265 | -0.123 | 0.902 | -2.634 | 2.323 |
|  | Group [MCI-R] | 1.875 | 1.364 | 1.375 | 0.169 | -0.798 | 4.548 |
|  | Group [MD] | 1.728 | 1.343 | 1.287 | 0.198 | -0.904 | 4.36 |
|  | Task Level [Detection 2] | 0.372 | 0.213 | 1.75 | 0.08 | -0.045 | 0.789 |
|  | Task Level [Resing State] | 0.551 | 0.308 | 1.793 | 0.073 | -0.051 | 1.154 |

Follow-up LMM analysis revealed that in the young healthy group (see table 6), VC9 showed increased activity for detection 2 compared to detection 1, and decreased activity for resting state (corrected *p*=0.012 *and p*=0.008 respectively). A0 activity significantly increased in detection 2 relative to detection 1 (corrected *p=*0.004). No significant differences between task levels were found in any of the senior groups (for all means and SE see figure 5 and 6, for LMM analyses variables see tables 5-7).

**Table 6.** Coefficients, Standard errors, z values, p values (before Bonferroni correction), and CI interval for task level (detection 1 as reference group) in the young healthy participants group.

| Feature | Variable [Level] | Coef. | Std.Err. | z | P>\|z\| | [0.025 | 0.975] |
| --- | --- | --- | --- | --- | --- | --- | --- |
| A0 | Intercept | 65.51 | 0.999 | 65.545 | <0.001 | 63.551 | 67.469 |
|  | Task Level [Detection 2] | 1.924 | 0.616 | 3.124 | **0.002** | 0.717 | 3.13 |
|  | Task Level [Resting State] | -0.283 | 0.787 | -0.36 | 0.719 | -1.825 | 1.259 |
| VC9 | Intercept | 52.589 | 0.971 | 54.149 | <0.001 | 50.685 | 54.492 |
|  | Task Level [Detection 2] | 2.161 | 0.794 | 2.722 | **0.006** | 0.605 | 3.717 |
|  | Task Level [Resting State] | -2.79 | 0.966 | -2.889 | **0.004** | -4.683 | -0.897 |
| Theta | Intercept | -6.4 | 0.592 | -10.817 | <0.001 | -7.559 | -5.24 |
|  | Task Level [Detection 2] | 1.119 | 0.424 | 2.635 | **0.008** | 0.287 | 1.951 |
|  | Task Level [Resting State] | -1.788 | 0.605 | -2.955 | **0.003** | -2.974 | -0.602 |
| Delta | Intercept | 5.38 | 0.641 | 8.396 | <0.001 | 4.124 | 6.636 |
|  | Task Level [Detection 2] | 1.417 | 0.558 | 2.54 | **0.011** | 0.324 | 2.51 |
|  | Task Level [Resting State] | -1.167 | 0.759 | -1.536 | 0.124 | -2.655 | 0.322 |

**Table 7.** Coefficients, Standard errors, z values, p values (before Bonferroni correction), and CI interval for group (healthy seniors as reference group), task level (detection 1 as reference group), and their interactions, in the senior groups (healthy seniors, MCI-R and MD).

| Feature | Variable [Level] | Coef. | Std.Err. | z | P>\|z\| | [0.025 | 0.975] |
| --- | --- | --- | --- | --- | --- | --- | --- |
| A0 | Task Level [Detection 2] | 1.4 | 0.892 | 1.57 | 0.116 | -0.347 | 3.148 |
|  | Task Level [Resting State] | 2.241 | 1.1 | 2.038 | **0.042** | 0.086 | 4.397 |
|  | Group [MCI-R] X Task Level [Detection 2] | -1.864 | 1.327 | -1.404 | 0.16 | -4.465 | 0.738 |
|  | Group [MD] X Task Level [Detection 2] | -0.846 | 1.293 | -0.655 | 0.513 | -3.381 | 1.688 |
|  | Group [MCI-R] X Task Level [Resting state] | -1.204 | 1.641 | -0.734 | 0.463 | -4.42 | 2.013 |
|  | Group [MD] X Task Level [Resting state] | -2.239 | 1.607 | -1.394 | 0.163 | -5.389 | 0.91 |
| VC9 | Task Level [Detection 2] | 0.38 | 0.478 | 0.795 | 0.427 | -0.557 | 1.316 |
|  | Task Level [Resting State] | -1.186 | 0.858 | -1.382 | 0.167 | -2.867 | 0.496 |
|  | Group [MCI-R] X Task Level [Detection 2] | 0.127 | 0.713 | 0.178 | 0.859 | -1.271 | 1.525 |
|  | Group [MD] X Task Level [Detection 2] | 0.764 | 0.688 | 1.111 | 0.267 | -0.584 | 2.111 |
|  | Group [MCI-R] X Task Level [Resting state] | -1.061 | 1.283 | -0.827 | 0.409 | -3.576 | 1.455 |
|  | Group [MD] X Task Level [Resting state] | 1.476 | 1.251 | 1.18 | 0.238 | -0.976 | 3.928 |
| Theta | Task Level [Detection 2] | 0.195 | 0.274 | 0.712 | 0.477 | -0.342 | 0.732 |
|  | Task Level [Resting State] | -0.546 | 0.513 | -1.065 | 0.287 | -1.552 | 0.46 |
|  | Group [MCI-R] X Task Level [Detection 2] | 0.229 | 0.408 | 0.561 | 0.575 | -0.571 | 1.029 |
|  | Group [MD] X Task Level [Detection 2] | 0.166 | 0.394 | 0.421 | 0.674 | -0.607 | 0.939 |
|  | Group [MCI-R] X Task Level [Resting state] | -0.576 | 0.763 | -0.755 | 0.45 | -2.071 | 0.919 |
|  | Group [MD] X Task Level [Resting state] | 0.621 | 0.749 | 0.829 | 0.407 | -0.847 | 2.089 |
| Delta | Task Level [Detection 2] | 0.414 | 0.391 | 1.058 | 0.29 | -0.353 | 1.181 |
|  | Task Level [Resting State] | -0.025 | 0.599 | -0.042 | 0.967 | -1.198 | 1.148 |
|  | Group [MCI-R] X Task Level [Detection 2] | -0.87 | 0.583 | -1.492 | 0.136 | -2.013 | 0.273 |
|  | Group [MD] X Task Level [Detection 2] | 0.583 | 0.563 | 1.036 | 0.3 | -0.52 | 1.687 |
|  | Group [MCI-R] X Task Level [Resting state] | -1.883 | 0.893 | -2.108 | **0.035\*** | -3.633 | -0.132 |
|  | Group [MD] X Task Level [Resting state] | 0.619 | 0.872 | 0.709 | 0.478 | -1.091 | 2.329 |

**Figure 5.**
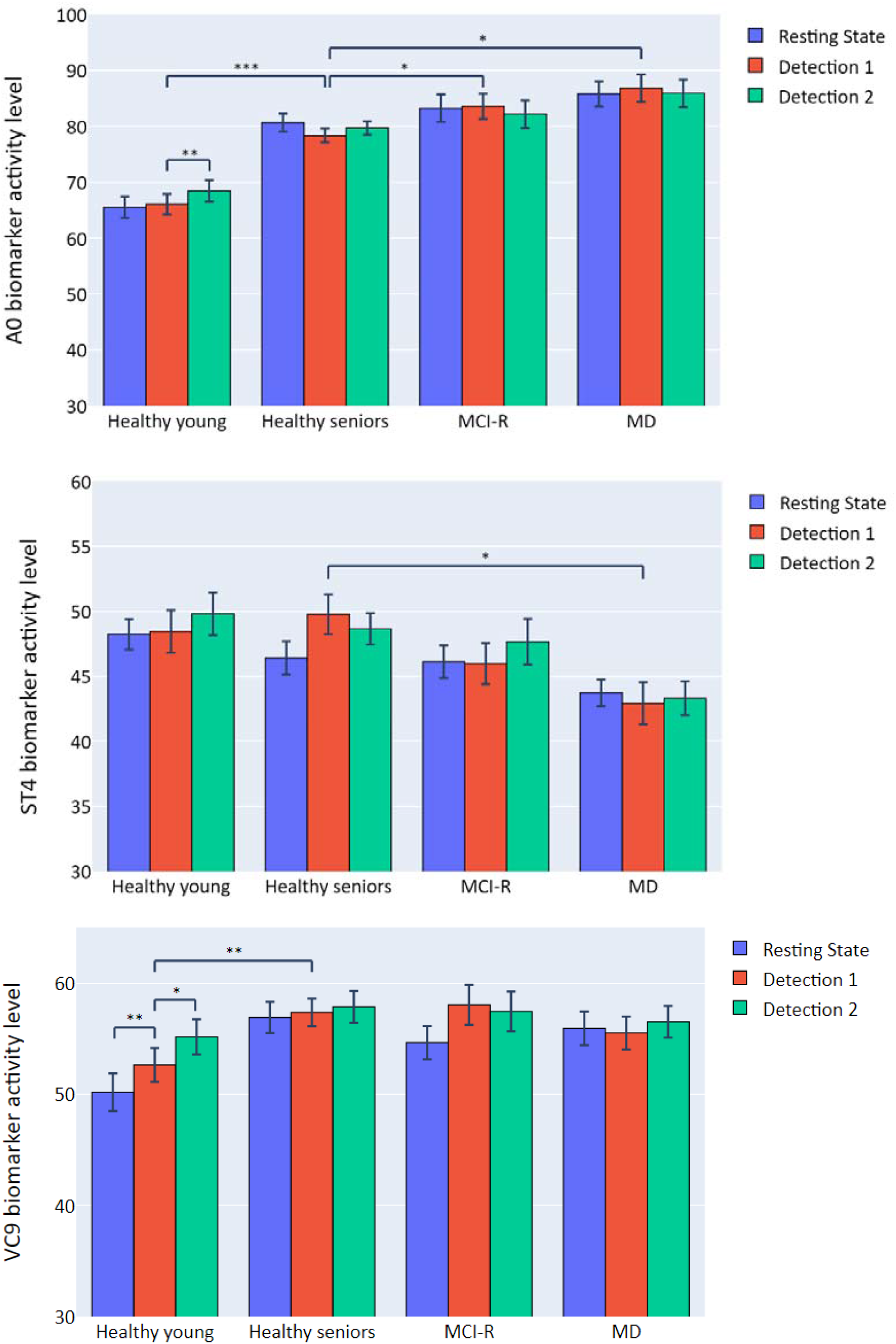
Mean activity of A0 (top), ST4 (middle), and VC9 (bottom) as a function of task: detection 2 (green), detection 1 (red) and resting state (blue), for the different groups. Data is presented as mean and SE.

**Figure 6.**
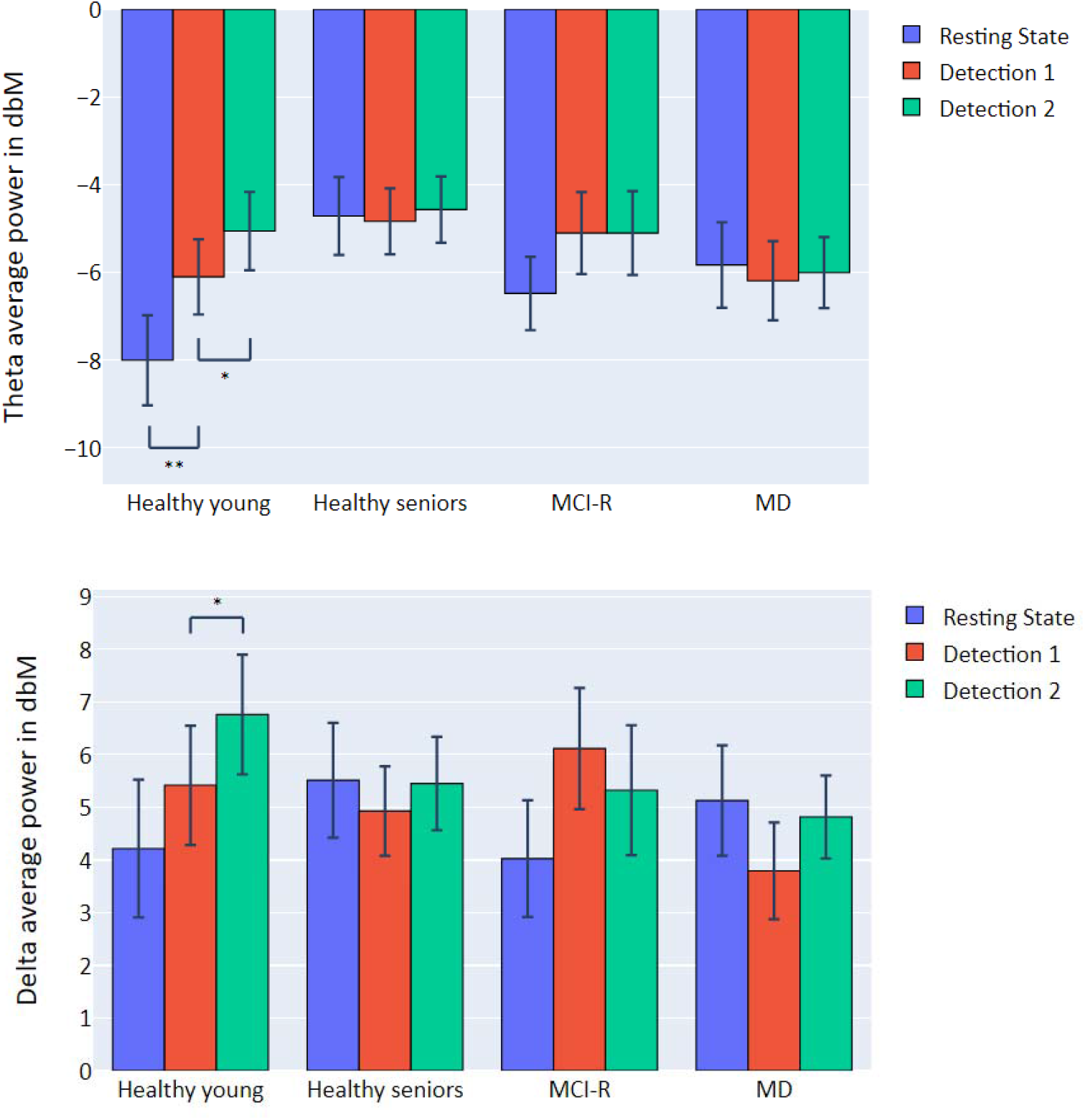
Mean activity of Theta (top) and Delta (bottom) as a function of task: detection 2 (green), detection 1 (red) and resting state (blue), for the different groups. Data is presented as mean and SE.

No significant differences were found between the groups in any of the frequency bands. However, Theta and Delta showed increased activity for detection 2 compared to detection 1 level (*p<*0.001, and *p=*0.006 respectively). Theta also exhibited a significant decrease in activity in resting state task compared to detection 1 (*p<*0.001).

Follow-up LMM analysis revealed that among the healthy young group, a significant increase in Theta activity in detection level 2 compared to detection level 1, as well as a significant decrease in resting state task (corrected *p*=0.006 *and p*=0.016 respectively). Delta activity in the healthy young group was significantly higher in detection level 2 relative to detection level 1 (corrected *p=*0.022). The difference in Delta power between detection 1 and resting state in the MCI-R group did not survive Bonferroni correction (corrected *p*=0.07). No other significant differences between task levels were found in any of the senior groups.

## Discussion

Cognitive decline remains highly underdiagnosed [28]. Improving the detection rate in the community to allow early intervention is therefore imperative. The aim of this study was to evaluate the ability of a single-channel EEG system with an interactive assessment tool to detect cognitive decline with correlation to known assessment methods. We demonstrate that a short evaluation which can be self-administered, together with automatically extracted objective EEG features from a wearable system with an easy set-up, may provide an assessment method for cognitive decline.

50 seniors and 22 young controls completed a short and interactive cognitive assessment battery. Classical EEG frequency bands were used in the analysis of the data as well as pre-defined Machine Learning (ML) features. ML applied on raw EEG signals is increasingly being examined for detection of cognitive deterioration. The biomarkers that are extracted using ML approaches show accurate separation between healthy and cognitively impaired populations [17], [18], [29]. Our approach utilizes advanced wavelet-packet analysis [30], [31] as pre-processing to ML. The biomarkers used here were calculated on a different dataset to avoid the risks associated with classification studies such as overfitting [32]. This is unlike other studies that use classifiers trained and tested via cross validation on the same dataset [10], [20], [21]. Specifically, the pre-extracted biomarkers used here, ST4 and VC9, were previously considered in a study performed on young healthy subjects. Results showed a correlation of VC9 to working memory load and correlation of ST4 to individual performance [23].

A novel interactive cognitive assessment based on auditory stimuli with three cognitive load levels (high, low and rest) was used to probe different cognitive states. Individual response performance in the tasks (i.e. reaction times) was correlated to the MMSE score which further validates the cognitive assessment tool. Importantly, individual response performance also significantly correlated with A0 and ST4 biomarkers. These results support previous findings that recording EEG during active engagement in cognitive and auditory tasks offer distinct features and may lead to better discrimination power of brain states [33]. To continue this notion, we used an auditory assessment battery with musical stimuli. It was previously shown that musical stimuli elicit stronger activity than using visual cues such as digits and characters [34].

Results further show that A0 and ST4 significantly correlated with individual MMSE scores. To get a clearer separation between cognitive stages, we divided the participants to groups according to the MMSE scores. In allocating the groups, we used the common cutoff score of 24 to divide between low-functioning (MMSE<24) and high-functioning participants, however, we divided the high-functioning group further using a cutoff score of 27, to get a notion of possible separability between cognitive states in high-functioning seniors.

Results demonstrated the ability of A0 and ST4 to significantly differentiate between groups of seniors with high vs. low MMSE scores. Specifically, ST4 separated between the healthy seniors and low-MMSE seniors (MD group) with the common cutoff score of 24, comparable to previous reports in this field [16], [20], [21]. Conversely, A0 showed differences between the healthy seniors and all other groups. A0 results suggest detection of more delicate differences between healthy seniors and seniors that are considered healthy to-date, but at a greater risk for developing cognitive decline (with MMSE scores below 27 but above 24). These results may indicate a different cognitive functionality between entirely healthy seniors and seniors that are not considered to suffer from cognitive decline, but score lower in the initial screening test, contributing to the debate in the literature over cognitive functionality of patients with scores below 27 [8], [26]. Furthermore, A0 and VC9 results suggest different cognitive patterns between healthy young participants and healthy seniors, consistent with reports from previous studies [35]. With regards to the commonly used EEG frequency bands, Theta and Delta activity increased with higher cognitive load, separating between the two cognitive load levels in the young healthy group. Theta also showed separation between low cognitive load and resting state. This is in line with previous reports of frontal Theta and Delta increase during cognitively demanding tasks [36]–[38]. This difference was not present in the senior population, supporting the notion that Theta and Delta may be indicative to cognitive decline and serve as a predictor of deterioration status, consistent with previous findings [10], [11]. Finally, A0 showed separation between high and low cognitive load levels within the cognitive task, further supporting the ability of A0 to differentiate between cognitive states. VC9 biomarker showed differences between all task levels, with a significant increase between rest and cognitive load levels within the cognitive task. This supports previous work validating VC9 as a working memory load biomarker in the healthy population [23]. Together, these results provide an initial indication of the ability of the proposed system to assess and detect cognitive decline in the elderly population.

It was shown that cognitive decline is associated with high-risk for developing dementia, and Alzheimer’s disease in particular [2], and that cognitive decline may be detected several years before dementia onset with known validated tools [39]. Dementia is gradually recognized as one of the most significant medical challenges of the future. So much so, that it has already reached epidemic proportions, with prevalence roughly doubling every five years over the age of 65 [40]. This rate is expected to increase unless therapeutic approaches are found to prevent or stop disease progression [41]. Since Alzheimer’s Disease (AD) is the most prevalent form of dementia, responsible for about 60–70% of cases [42], it remains the focus of clinical trials. To date, most clinical trials that include a disease-modifying treatment, fail to demonstrate clinical benefits in symptomatic AD patients. This could be explained by the late intervention that occurs after neuropathological processes have already resulted in substantial brain damage [43]. Hence, the discovery of predictive biomarkers for preclinical or early clinical stages such as cognitive decline is imperative [44].

The ability to differentiate between cognitive sates with the novel biomarkers shown here relies only on a single EEG channel and a short assessment battery, unlike most studies attempting to assess cognitive decline with multichannel EEG systems [17], [45], [46]. It has been argued that the long setup time of multichannel EEG systems may cause fatigue, stress, or even change mental states, affecting EEG patterns and subsequently study outcomes [22]. This suggests that cognitive decline evaluation using a wearable single-channel EEG with a quick setup time may not only make the assessment more affordable and accessible but also potentially reduce the effects of the pretest time on the results. Using a single EEG channel was previously shown to be effective in detection of cognitive decline [20], however, here we demonstrate results using biomarkers that were extracted on an independent dataset to avoid an overfitting of the data. The extracted cognitive biomarkers from the system offered here may potentially enable detection in the community in much earlier stages before dementia symptoms arise.

While this pilot study shows initial promising results, more work is needed. Specifically, additional studies should include a longer testing period to quantify the variability within subjects and potentially increase the predictive power. Also, larger cohorts of patients that are quantified by extensive screening methods would offer an opportunity to get more sensitive separation between earlier stages of cognitive decline using the suggested tool, and potentially reduce the subjective nature of the MMSE. Moreover, a longitudinal study could assess cognitive state in asymptomatic elderly patients and follow participants’ cognition over an extended period of time, validating the predictive power of the biomarkers.

## Conclusions

In conclusion, this pilot study successfully demonstrated the ability to assess cognitive decline, using a wearable single-channel EEG system and novel cognitive biomarkers which correlate to well-validated clinical measurements for detection of cognitive decline. Using a low-cost approach and objective biomarkers, it may provide consistency in assessment across patients and between medical facilities clear of tester bias. Furthermore, due to a short set-up time and interactive cloud-based assessment, this tool has the potential to be used on a large scale in every clinic to detect deterioration before clinical symptoms emerge. Future studies should explore potential usefulness of this tool in characterizing changes in EEG patterns of cognitive decline over time, for early detection of cognitive decline to potentially allow earlier intervention.

## Data Availability

The data that support the findings of this study are available on request from the corresponding author. The data are not publicly available due to restrictions (containing information that could compromise the privacy of research participants).

## Acknowledgements/Conflicts/Funding Sources

The authors wish to thank Talya Zeimer for her support while writing this manuscript. We thank the study participants and the supporting staff for their contributions to this study.

L.M., N.B.M. and N.I. have equity interest in Neurosteer, which developed the Neurosteer Aurora system. The other authors have no competing interests to declare.

## Notes

### Competing Interest Statement

The authors have declared no competing interest.

### Clinical Trial

NCT04386902

### Funding Statement

No external funding was received for this study. The equipment was provided by Neurosteer Inc. as part of the trial.

### Author Declarations

Ethical approval for this study was granted by the Ethics Committee (EC) of Dorot Geriatric Medical Center.

### Summary of Updates

Data analysis section updated with LMM analysis; Results and discussion updated accordingly. Figure 5-6 revised; Tables 5-7 revised.

## References

[1] S. Plassman, B.L., Williams Jr, J.W.Burke, J. R., Holsinger, T., & Benjamin, “Systematic review: factors associated with risk for and possible prevention of cognitive decline in later life.,” Ann. Intern. Med., vol. 153, no. 3, pp. 182–193, 2010, doi: 10.7326/0003-4819-153-3-201008030-00258.

[2] K. Ritchie and J. Touchon, “Mild cognitive impairment: Conceptual basis and current nosological status,” Lancet. 2000, doi: 10.1016/S0140-6736(99)06155-3.

[3] N. B. Silverberg et al., “Assessment of cognition in early dementia,” Alzheimer’s Dement., 2011, doi: 10.1016/j.jalz.2011.05.001.

[4] C. B. Cordell et al., “Alzheimer’s Association recommendations for operationalizing the detection of cognitive impairment during the Medicare Annual Wellness Visit in a primary care setting,” Alzheimer’s and Dementia. 2013, doi: 10.1016/j.jalz.2012.09.011.

[5] M. F. Folstein, S. E. Folstein, and P. R. McHugh, “‘Mini-mental state’. A practical method for grading the cognitive state of patients for the clinician,” J. Psychiatr. Res., 1975, doi: 10.1016/0022-3956(75)90026-6.

[6] T. Tombaugh and N. McIntyre, “Tombaugh 1992 The MMSE - A comprehensive review.pdf,” Journal of the American Geriatrics Society. 1992.

[7] R. M. Crum, J. C. Anthony, S. S. Bassett, and M. F. Folstein, “Population-Based Norms for the Mini-Mental State Examination by Age and Educational Level,” JAMA J. Am. Med. Assoc., 1993, doi: 10.1001/jama.1993.03500180078038.

[8] J. S. Shiroky, H. M. Schipper, H. Bergman, and H. Chertkow, “Can you have dementia with an MMSE score of 30?,” Am. J. Alzheimers. Dis. Other Demen., 2007, doi: 10.1177/1533317507304744.

[9] R. Cassani, M. Estarellas, R. San-Martin, F. J. Fraga, and T. H. Falk, “Systematic review on resting-state EEG for Alzheimer’s disease diagnosis and progression assessment,” Disease Markers. 2018, doi: 10.1155/2018/5174815.

[10] F. R. Farina et al., “A comparison of resting state EEG and structural MRI for classifying Alzheimer’s disease and mild cognitive impairment,” Neuroimage, 2020, doi: 10.1016/j.neuroimage.2020.116795.

[11] D. M.P. et al., “Attention and Working Memory-Related EEG Markers of Subtle Cognitive Deterioration in Healthy Elderly Individuals,” J. Alzheimer’s Dis., 2015.

[12] P. Missonnier et al., “Working memory load-related electroencephalographic parameters can differentiate progressive from stable mild cognitive impairment,” Neuroscience, 2007, doi: 10.1016/j.neuroscience.2007.09.009.

[13] J. Dauwels, F. Vialatte, and A. Cichocki, “Diagnosis of Alzheimer’s Disease from EEG Signals: Where Are We Standing?,” Curr. Alzheimer Res., 2010, doi: 10.2174/1567210204558652050.

[14] W. S. Pritchard et al., “EEG-based, neural-net predictive classification of Alzheimer’s disease versus control subjects is augmented by non-linear EEG measures,” Electroencephalogr. Clin. Neurophysiol., 1994, doi: 10.1016/0013-4694(94)90033-7.

[15] C. Babiloni et al., “Mapping distributed sources of cortical rhythms in mild Alzheimer’s disease. A multicentric EEG study,” Neuroimage, 2004, doi: 10.1016/j.neuroimage.2003.09.028.

[16] C. Lehmann et al., “Application and comparison of classification algorithms for recognition of Alzheimer’s disease in electrical brain activity (EEG),” J. Neurosci. Methods, 2007, doi: 10.1016/j.jneumeth.2006.10.023.

[17] A. Cichocki, S. L. Shishkin, T. Musha, Z. Leonowicz, T. Asada, and T. Kurachi, “EEG filtering based on blind source separation (BSS) for early detection of Alzheimer’s disease,” Clin. Neurophysiol., 2005, doi: 10.1016/j.clinph.2004.09.017.

[18] C. Melissant, A. Ypma, E. E. E. Frietman, and C. J. Stam, “A method for detection of Alzheimer’s disease using ICA-enhanced EEG measurements,” Artif. Intell. Med., 2005, doi: 10.1016/j.artmed.2004.07.003.

[19] M. Ahmadlou, H. Adeli, and A. Adeli, “New diagnostic EEG markers of the Alzheimer’s disease using visibility graph,” J. Neural Transm., 2010, doi: 10.1007/s00702-010-0450-3.

[20] S. Khatun, B. I. Morshed, and G. M. Bidelman, “A Single-channel EEG-based approach to detect mild cognitive impairment via speech-evoked brain responses,” IEEE Trans. Neural Syst. Rehabil. Eng., 2019, doi: 10.1109/TNSRE.2019.2911970.

[21] M. Kashefpoor, H. Rabbani, and M. Barekatain, “Automatic diagnosis of mild cognitive impairment using electroencephalogram spectral features,” J. Med. Signals Sens., 2016, doi: 10.4103/2228-7477.175869.

[22] R. Cassani, T. H. Falk, F. J. Fraga, M. Cecchi, D. K. Moore, and R. Anghinah, “Towards automated electroencephalography-based Alzheimer’s disease diagnosis using portable low-density devices,” Biomed. Signal Process. Control, 2017, doi: 10.1016/j.bspc.2016.12.009.

[23] N. B. Maimon, L. Molcho, N. Intrator, and D. Lamy, “Single-channel EEG features during n-back task correlate with working memory load,” Aug. 2020, Accessed: Oct. 06, 2020. [Online]. Available: http://arxiv.org/abs/2008.04987.

[24] R. R. Coifman and M. V. Wickerhauser, “Entropy-based algorithms for best basis selection,” IEEE Trans. Inf. Theory, 1992, doi: 10.1109/18.119732.

[25] N. Neretti and N. Intrator, “An adaptive approach to wavelet filters design,” 2002, doi: 10.1109/NNSP.2002.1030043.

[26] S.E. O’Bryant et al., “Detecting dementia with the mini-mental state examination in highly educated individuals,” Arch. Neurol., 2008, doi: 10.1001/archneur.65.7.963.

[27] J. Seabold S., & Perktold, “Statsmodels: Econometric and statistical modeling with python,” in In Proceedings of the 9th Python in Science Conference, p. Vol. 57, 61.

[28] L. Lang et al., “Prevalence and determinants of undetected dementia in the community: A systematic literature review and a meta-analysis,” BMJ Open. 2017, doi: 10.1136/bmjopen-2016-011146.

[29] J. P. Amezquita-Sanchez, N. Mammone, F. C. Morabito, S. Marino, and H. Adeli, “A novel methodology for automated differential diagnosis of mild cognitive impairment and the Alzheimer’s disease using EEG signals,” J. Neurosci. Methods, 2019, doi: 10.1016/j.jneumeth.2019.04.013.

[30] N. Intrator, “Systems and methods for brain activity interpretation,” USA Patent US20160235351A1, 2018.

[31] N. Intrator, “Systems and methods for analyzing brain activity and applications thereof,” USA Patent US20170347906A1, 2019.

[32] M.-P. J.M., D. M., L.-A. M., I.-M. Y., Z. Y., and E. A.C., “Structural neuroimaging as clinical predictor: A review of machine learning applications,” NeuroImage Clin., 2018.

[33] P. Ghorbanian et al., “Identification of resting and active state EEG features of alzheimer’s disease using discrete wavelet transform,” Ann. Biomed. Eng., 2013, doi: 10.1007/s10439-013-0795-5.

[34] M. Tervaniemi, A. Kujala, K. Alho, J. Virtanen, R. J. Ilmoniemi, and R. Näätänen, “Functional specialization of the human auditory cortex in processing phonetic and musical sounds: A magnetoencephalographic (MEG) study,” Neuroimage, 1999, doi: 10.1006/nimg.1999.0405.

[35] I. et al. Vlahou, E., Thurm, F., Kolassa, “Resting-state slow wave power, healthy aging and cognitive performance,” Sci Rep, vol. 4, p. 5101, 2014, doi: https://doi.org/10.1038/srep05101.

[36] R.M. & A. D. N. I. Hadjichrysanthou, C. Evans, S., Bajaj, S., Siakallis, L. C., McRae-McKee, K., de Wolf, F., Anderson, “The dynamics of biomarkers across the clinical spectrum of Alzheimer’s disease,” Alzheimers. Res. Ther., vol. 12, no. 1, p. 74, 2020, doi: https://doi.org/10.1186/s13195-020-00636-z.

[37] W. M. Van Der Flier and P. Scheltens, “Epidemiology and risk factors of dementia,” Neurology in Practice. 2005, doi: 10.1136/jnnp.2005.082867.

[38] L. E. Hebert, J. Weuve, P. A. Scherr, and D. A. Evans, “Alzheimer disease in the United States (2010-2050) estimated using the 2010 census,” Neurology, 2013, doi: 10.1212/WNL.0b013e31828726f5.

[39] Q. C., K. M., and V. S. E., “Epidemiology of Alzheimer’s disease: Occurrence, determinants, and strategies toward intervention,” Dialogues Clin. Neurosci., 2009.

[40] D. Galimberti and E. Scarpini, “Disease-modifying treatments for Alzheimer’s disease,” Therapeutic Advances in Neurological Disorders. 2011, doi: 10.1177/1756285611404470.

[41] C. R. J. Jr et al., “Introduction to revised criteria for the diagnosis of Alzheimer’s disease: National Institute on Aging and the Alzheimer Association Workgroups,” Alzheimer Dement., 2011, doi: 10.1016/j.jalz.2011.03.004.Introduction.

[42] J. Dauwels, F. Vialatte, T. Musha, and A. Cichocki, “A comparative study of synchrony measures for the early diagnosis of Alzheimer’s disease based on EEG,” Neuroimage, 2010, doi: 10.1016/j.neuroimage.2009.06.056.

[43] D. V. Moretti et al., “MCI patients’ EEGs show group differences between those who progress and those who do not progress to AD,” Neurobiol. Aging, 2011, doi: 10.1016/j.neurobiolaging.2009.04.003.

[44] O. Jensen and C. D. Tesche, “Frontal theta activity in humans increases with memory load in a working memory task,” Eur. J. Neurosci., 2002, doi: 10.1046/j.1460-9568.2002.01975.x.

[45] R. Scheeringa, K. M. Petersson, R. Oostenveld, D. G. Norris, P. Hagoort, and M. C. M. Bastiaansen, “Trial-by-trial coupling between EEG and BOLD identifies networks related to alpha and theta EEG power increases during working memory maintenance,” Neuroimage, 2009, doi: 10.1016/j.neuroimage.2008.08.041.

[46] T. Harmony and T. Farnandez, “EEG delta activity: an indicator of attention to internal processing during performance of mental tasks,” Int. J. Psychophysiol., pp. 161–171, 1996.

[47] F. P. Zarjam, J. Epps, & Chen, “Characterizing working memory load using EEG delta activity,” in 19th Eusipco Conference, 2011, pp. 1554–1558.

[48] I. Schapkin, S.A. Raggatz, J., Hillmert, M., & Böckelmann, “EEG correlates of cognitive load in a multiple choice reaction task,” Acta Neurobiol., vol. 80, pp. 76–89, 2020.

