## Appendix A for "Detection of cognitive decline using a single-channel EEG with an interactive assessment tool"

### Appendix A: Methodological details

The data analysis methodology is composed of three steps:

1. Creation of a brain activity representation by novel Brain Activity Features (BAFs)
2. Creation of Novel Biomarkers based on the BAFs
3. Examination of the features on previously unseen data

Each of the steps is described below.

*Creation of Brain Activity features (BAFs)*

The creation of the Brain Activity Features (BAFs) occurs prior to application of the methodology onto the new data to be analyzed. Calculation of the BAFs is based on collecting a large cohort of high dynamic amplitude and frequency range single channel EEG data. The cohort includes multiple subjects that are exposed to different cognitive, emotional, and resting tasks. A schematic representation of the signal processing is depicted in Fig A1. The signal processing module is decomposing the EEG signal input into a large number of components which comprise the Brain Activity Features (BAFs). The output of the module is a Brain Activity Representation which is constructed based on the BAFs for any given EEG signal.

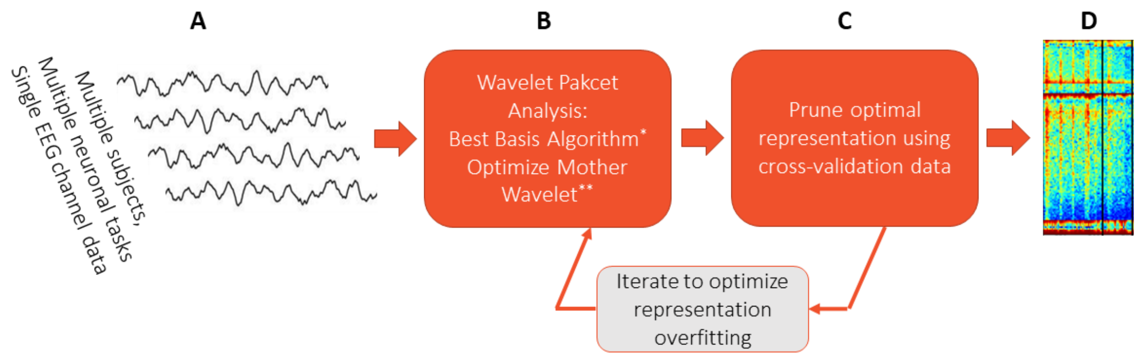

Figure A1. schematic representation of the construction of the Brain Activity Features (BAFs). See text for the description of the different steps.

**A: electrophysiological signal input**

The EEG cohort described above is the input of the signal processing algorithm presented as the first step of the process.

**B: Wavelet Packet Analysis**

For a given cohort of EEG recordings, a family of *wavelet packet trees* is created. For the mathematical description, we follow the notation and construction provided in chapters 5, 6 and 7 of Wickerhauser's book[1].

To demonstrate the process; let *g* and *h* be a set of *biorthogonal quadrature filters* created from the filters *G* and *H* respectively. Each of these is a convolution-decimation operator, where in the case of the simple *Haar* wavelet, *g* is a set of averages and *h* is a set of differences.

The construction of the full wavelet packet tree is by successive application of these functions (Figure A2), so that at every level, a new full orthogonal decomposition of the original signal *x* is created. In the classical wavelet decomposition by Daubechies[2], only the marked parts are used and the signal is decomposed into *Gx, GHx etc.,* but the full construction of the tree continues recursively, on *Gx, GHx* and so forth, to create a full binary tree. Coifman and Wickerhauser[3] observed that a large number of orthogonal decompositions can be constructed from the full tree by mixing between the different levels and different blocks of the tree, following a simple rule. The recursive construction of the full tree is described next.

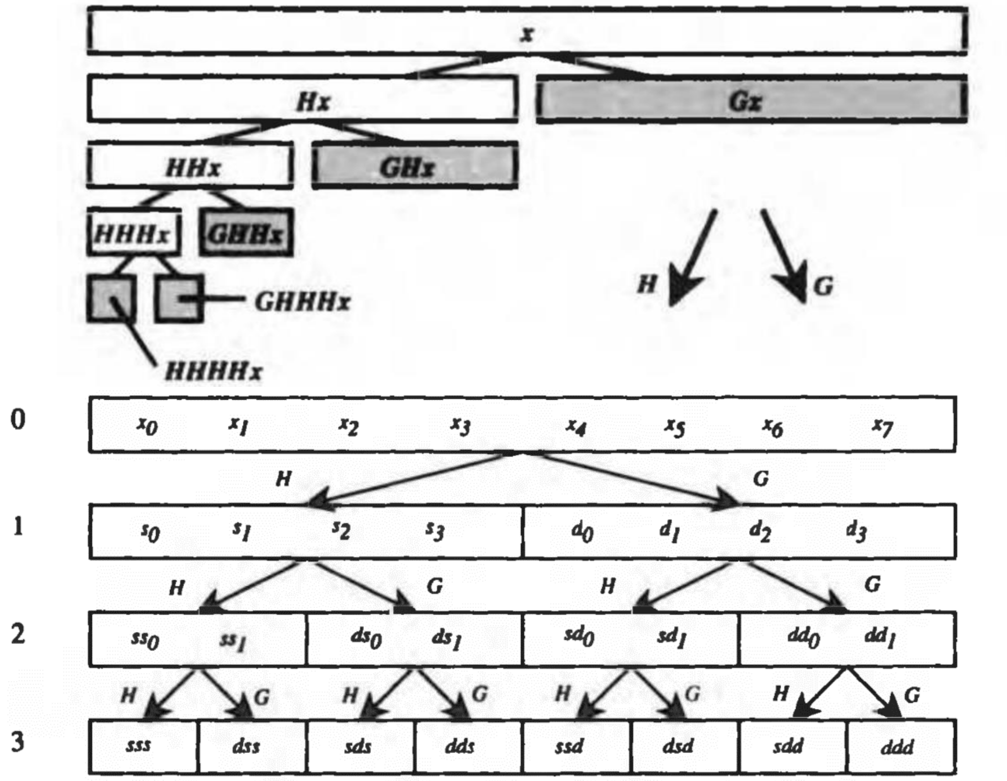


Figure A2. Construction of a Discrete Wavelet Transform Tree (Taken from Wickerhauser[1]). The top panel represents the classical wavelet construction and the bottom panel extends the construction to a full wavelet packet tree.

Let $\psi_1$ be the *mother wavelet* associated to the filters $s \in H$, an $d \in G$. Then, the collection of *wavelet packets* $\psi_n$, is given by:

$$\psi_{2n} = H\psi_n; \qquad \psi_{2n}(t) = \sqrt{2}\sum_{j\in Z} s(j)\psi_n(2t-j),$$

$$\psi_{2n+1} = G\psi_n; \qquad \psi_{2n+1}(t) = \sqrt{2}\sum_{j\in Z} d(j)\psi_n(2t-j).$$

The recursive form provides a natural arrangement in the form of a binary tree (Figure A2). The functions $\psi_n$ have a fixed scale. A library of wavelet packets of any scale *s*, frequency *f*, and position *p* is given by:

$$\psi_{sfp}(t) = 2^{-s/2}\psi_f(2^{-s}t - p).$$

The wavelet packets $\{\psi_{sfp}: p \ \in Z\}$ are an orthonormal basis for every $f$ (under orthogonality condition of the filters $H$ and $G$) and are called $orthonomal\ wavelet\ packets$.

Using this construction, Coifman and Wickerhauser applied the *best basis* algorithm[3] to search for an orthonormal base that satisfies a specific optimality condition. The optimality condition that was chosen is Shannon's entropy of the coefficients of each component (or wavelet packet atom). It is a measure that prefers coefficients with a distribution that is far from uniform, in the sense that it prefers a distribution with a small number of high value coefficients and a long tale, namely, a large number with low value coefficients. The full details of the best basis search are described in chapter 7 of Wickerhaser's book.

The process of creating a best basis from the wavelet packet tree can be further iterated by an optimization on the mother wavelet using a gradient descent in wavelet space as is described in Neretti and Intrator[4].

**C: Pruning the optimal representation**

The outcome of the best basis algorithm is an orthogonal decomposition that is adapted to the stochastic properties of the collection of EEG signals. However, there is a risk that the decomposition is "overfitting" namely it is too adapted to the EEG signals from which it was created. To avoid this phenomenon, we first have to get rid of "small" coefficients. This can be done by the denoising technique of Coifman and Donoho[5]. The next step is introducing a validation set, which is another collection of EEG-recordings that was not used in the creation of the best basis. Using this set, we can determine which atoms maintain a high energy (some large coefficients) when decomposing the new signals. These atoms will remain in the representation. At the end of this part, the resulting set of decomposing signal contains only a part of the full orthonormal basis that was found. We then reorder the basis components not based on the binary tree that created them, but based on the correlation between the different components In this way, we created a brain activity representation in which components that are more correlated to each other, are also geographically close to each other within the representation. This is done for the purpose of improved visualization.

**D: brain activity representation output**

The result of the signal processing module is the brain activity representation. Specifically, it is a collection of 121 energy components, emanating from the wavelet packets as well as standard frequency bands which are updated each second. The representation (D) shows a color heatmap of each of the 121 X time matrix, so that the x axis represents time and the y axis represents the different components.

*Creation of Novel features based on the BAFs*

The signal components, which we termed BAFs, were constructed from single EEG channel recordings in an unsupervised manner, namely, there were no labels attached to the recordings for the purpose of creating the decomposition. To create biomarkers based on the BAFs, task labels are used, indicating the nature of cognitive, emotional, or resting challenge the subject is exposed to during the recording.

Given labels from a collection of subjects, and the corresponding high-dimensional BAF data, a collection of models attempting to differentiate between the labels based on the BAF activity can be used. In the linear case, these models are of the form:

$$V_k(w, x) = \Psi\left(\sum_i w_i x_i\right),$$

where w is a vector of weights, and $\Psi$ is a transfer function that can either be linear, e.g., $\Psi(y) = y$, or sigmoidal for logistic regression $\Psi(y) = 1/(1 + e^{-y})$.

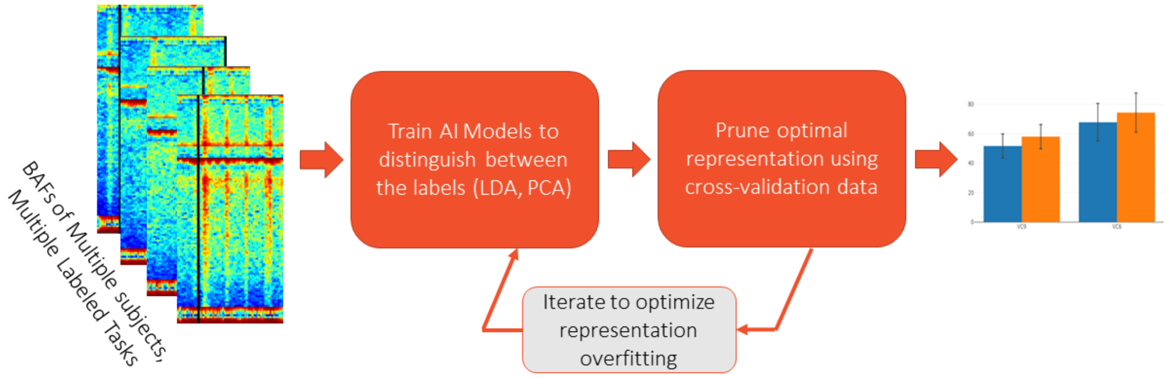


Figure A3. Supervised construction of different features from labeled brain activity representation of different cognitive and non-cognitive tasks.

For each predictor, which we term biomarker, a standard machine learning procedure is applied as follows:

1. Choose a labeled data set with at least two different tasks (e.g. cognitive, emotional, or resting challenge). The data set may include the same challenge but for a non-homogenous group.
2. Separate the data into three sets: training, validation, and test.
3. Choose a model to train on from a family of models that includes linear regression, linear regression with binary constrains (zero and one values for the weights), linear regression with only positive values, logistic regression, discriminant analysis and principal components analysis. In the non-linear models, use neural networks, support vector machine and the like.

4. Train each model on several sets of train/test and validation to best estimate internal model such as the variance constraints, on the ridge regression, the kernel size and number of kernels in a support vector machine, or the weight constraints in a neural network model.
5. From the above models, obtain predictors to be tested on other data with potentially other cognitive, emotional and rest challenges.
6. The last step in the process includes testing the biomarkers using a test data labeled set that was not used in the creation of these features. This allows removal of features that were overfitting to the training data, namely, they do not produce high significant difference on the validation data. This is still part of the model creation and not part of the model testing that is done on new data and is described in step 3.

All above steps are described in the scheme on Figure A3.

*Examination of the features on previously unseen data*

Following the creation of BAFs and the creation of features as described above, the features relevance can be tested on various cognitive or emotional challenge. The testing scheme is described in Figure A4. Specifically, data is collected with the sensor system and sent to the cloud for creation of a BAF representation using the previously determined wavelet packet atoms. The BAF representation is provided to previously determined ML models, which convert the BAF activity into features. Statistical tests are then applied to determine the quality of the predictions and the correlation of the features to the cognitive and emotional challenges that the participants undergo. This may include single subject analysis as well as group analysis.

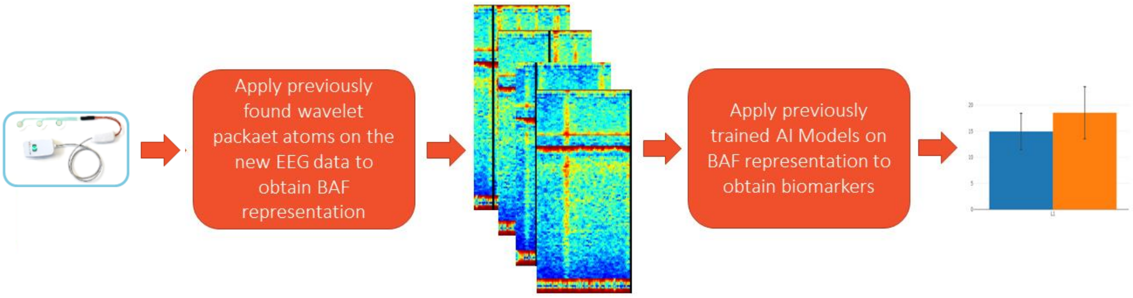


Figure A4: Testing the relevance of the previously found features on the data.

In the process of testing the features on new data, we may want to get an *upper bound* to the performance of the feature, by seeking an *overfitting biomarker* on the currently tested data. This is only done to get an idea of the potential upper bound on prediction abilities from the existing data, and indirectly can tell us more about the optimality of the actual features that were constructed from a different data set and are assumed to be more general in this sense.

#### Appendix References


[1] Wickerhauser, M. V. (1996). Adapted wavelet analysis: from theory to software. CRC Press.

[2] Daubechies, Ingrid. (1988). "Orthonormal bases of compactly supported wavelets." Communications on pure and applied mathematics, 41(7): 909-996.

[3] Coifman, R.R., and M.V. Wickerhauser. (1992). “Entropy-Based Algorithms for Best Basis Selection.” IEEE Transactions on Information Theory, 38(2): 713-718.

[4] Neretti N, Intrator N. (2002). An adaptive approach to wavelet filters design. Neural Networks for Signal Processing. Proceedings of the 2002 12$^{th}$ IEEE Workshop on IEEE.

[5] Coifman R.R. and D. L. Donoho. (1995). “Translation-Invariant De-Noising.” Wavelets and Statistics, 125-150.
